# Use of portable air cleaners to reduce aerosol transmission on a hospital COVID-19 ward

**DOI:** 10.1101/2021.03.29.21254590

**Authors:** KL Buising, R Schofield, L Irving, M Keywood, A Stevens, N Keogh, G Skidmore, I Wadlow, K Kevin, B Rismanchi, AJ Wheeler, RS Humphries, M Kainer, F McGain, J Monty, C Marshall

## Abstract

**Objective:** To study the airflow, transmission and clearance of aerosols in the clinical spaces of a hospital ward that had been used to care for patients with COVID-19, and to examine the impact of portable air cleaners on aerosol clearance.

**Design:** Observational study

**Setting:** A single ward of a tertiary public hospital in Melbourne Australia

**Intervention:** Glycerine-based aerosol was used as a surrogate for respiratory aerosols. The transmission of aerosols from a single patient room into corridors and a nurses’ station in the ward was measured. The rate of clearance of aerosols was measured over time from the patient room, nurses’ station and ward corridors with and without air cleaners (also called portable HEPA filters).

**Results:** Aerosols rapidly travelled from the patient room into other parts of the ward. Air cleaners were effective in increasing the clearance of aerosols from the air in clinical spaces and reducing their spread to other areas. With two small domestic air cleaners in a single patient room of a hospital ward, 99% of aerosols could be cleared within 5.5 minutes.

**Conclusion:** Air cleaners may be useful in clinical spaces to help reduce the risk of healthcare acquired acquisition of respiratory viruses that are transmitted via aerosols. They are easy to deploy and are likely to be cost effective in a variety of healthcare settings

---

COVID-19 presents a major global health challenge, with extraordinary clinical, societal and economic impacts. While initially there may have been controversy on the role of airborne transmission, now most authorities suggest that transmission of SARS-CoV-2 (the virus causing COVID-19) can occur via contact, droplet and/or airborne routes depending on the circumstances.^1-8^ The different mechanisms of transmission likely arise due to variation in the size of respiratory particles generated, which depends on a number of factors including, but not limited to, the site of primary infection (lower vs upper respiratory tract), exposure to aerosol generating procedures (e.g.; nebulizer use) or aerosol generating behaviours of the source (e.g.; coughing, shouting, singing).^9-16^ The context is also likely to be very important, for example, airborne transmission may be more likely to occur where there are several infected people (sources) in a confined space with a limited clearance of aerosolised particles due to poor ventilation.^17-19^

In many countries, an increased risk of infection with SARS-CoV-2 has been reported amongst frontline healthcare workers. At the Royal Melbourne Hospital, 271 healthcare workers acquired COVID-19 infection, and the consequent epidemiologic and genomic investigation suggested that most of these were healthcare acquired.^20^ The vast majority of these infections seemed to occur in wards that were designated to care for patients already known to be infected, where staff were already using infection prevention precautions appropriate to the risk and recommendations at the time. Clinicians observed that transmission to healthcare workers seemed to be more common when the number of concurrent COVID-19 infected patients in a given ward was high, and in association with particular behaviours (e.g.; shouting, patients with high respiratory rate or persistent cough). Staff without patient contact were also noted to become infected, especially if they spent prolonged times in the ward (namely corridors and nurses’ stations), but not necessarily in the patient rooms. The high rate of staff infection raised concern about possible aerosol transmission, so staff within the wards were required to wear N95/P2 masks rather than surgical masks in mid-July ahead of state and national guidance. A group of multidisciplinary researchers from engineering, aerosol science, virology, infection prevention, and infectious diseases and respiratory medicine was convened to discuss what was known about aerosol transmission of SARS-CoV-2 in July 2020 and met weekly thereafter. As new evidence emerged to support that hypothesis, the focus quickly moved to considering what mitigation strategies could be used in a clinical space to reduce risks for staff.^21-29^ Information about the efficacy of air cleaners, tested in laboratory conditions was shared and the group decided that work was needed to understand the effects of directional airflow in clinical spaces that might help explain local infection observations. In addition, improved understanding of the utility of air cleaners in ward environments was needed to help inform hospital policy decisions. This study aimed to trace airflow within a ward where known COVID-19 patients had been cared for and in common spaces outside the ward, and to document the effectiveness of air cleaners in reducing airborne particle concentrations.

## Setting

The Royal Melbourne Hospital is a university affiliated tertiary hospital with 550 acute and 150 subacute (rehabilitation, geriatric medicine) inpatient beds. The hospital has cared for the largest number of inpatients with COVID-19 in Australia to date, namely 525 inpatients, with a peak of 99 concurrent inpatients, predominantly in July and August 2020 coinciding with the second wave of the pandemic in Victoria. The infectious diseases ward has 14 negative pressure rooms with anterooms, along with negative pressure rooms in the emergency department, acute medical assessment unit and the intensive care unit. The negative pressure rooms conform to the relevant Australian standards as Class N/Type 5 rooms. As the number of concurrent inpatients with COVID-19 infection climbed and the capacity of the infectious diseases ward was exceeded, five other medical and surgical wards were progressively converted to “COVID wards”. Staff wore long sleeved gowns, gloves, eye protection (full face shield) and N95/P2 mask when looking after patients. All staff entering the ward were required to wear PPE in rooms, corridors and nurses’ stations.

In December 2020, one of the wards that had been used for patients with COVID-19 during July and August 2020 was vacated for 2 weeks due to planned low surgical activity over the Christmas holiday period. The research team had full access to the empty ward to conduct in-situ experiments assessing airflow and aerosol clearance over this time. This ward has a long central corridor and 11 rooms, which usually accommodate 25 patient beds (four single rooms with own ensuite bathroom and seven three-bed shared rooms each with a shared ensuite bathroom). The nurses’ station is situated halfway along the corridor, directly opposite two single rooms. No rooms in this ward have negative pressure, and the ward has its own closed, ducted heating, ventilation and air conditioning (HVAC) system delivering 12 air changes per hour. No windows in the ward can be opened, and the return air vent for the whole ward is above the single entrance and exit point to the ward (just inside the door to the ward). Rooms all have doors, which have a small gap at the bottom (approximately 5cm) to allow air egress.

Many patients with COVID-19 who were cared for in that ward during the peak of the pandemic (July and August) were residents from aged care homes. Many were elderly and required high levels of nursing care. Patients were predominantly cared for with one patient per room, but at times two patients were managed in a space designed for three-beds. The peak concurrent number of patients in the ward was 15. Due to frailty, delirium and fall risk, the doors to patient rooms sometimes needed to stay open to provide appropriate supervision and nursing care.

## Methods

One of the single patient rooms with a room floor space of 12.8 m^2^ and volume of approximately 37 m^3^ was selected for the study. The corridor outside the room was approximately 2 m wide, and the patient room was directly opposite the open nurses’ station, which had a front desk with entrance spaces on either side. For each experiment, glycerine-based aerosol (smoke) with a mean aerosol size of 1 µm was injected for 15 seconds into the patient room to ‘flood’ the air space with ‘smoke’. The particle sizes of this smoke roughly approximate the sizes of SARS-CoV-2 infectious aerosols measured in the air of hospitals elsewhere (<1 µm) (Brigand et al., 2020).^30^ We used glycerine-based aerosol as a surrogate method to replicate aerosol movement and airborne transmission. Once injected, it was permitted to mix and visibly fill all corners of the room. The visible pathways of travel of the smoke within the patient room, corridor and nurses’ station were observed and video recorded.

The sensors used in this experiment to measure aerosols were the TSI DustTrak DRX 8533, (called a “DRX”), and the TSI DustTrak II 8530, (called a “DRII”). The DustTrak sensors measured aerosol concentration using a combination of particle cloud and single particle detection to measure the mass concentration of aerosols per unit volume. The DustTraks sensors were fitted with a 2.5 µm inlet that only allowed aerosols 2.5 µm or smaller to pass through. The DRII was placed inside the patient room (or corridor) and the DRX was placed in the nurses’ station to determine the amount of aerosol that could move through the corridor or across into the nurses’ station.

Measurements were taken concurrently in the single patient room, and at the nurses’ station at 10-second intervals until the aerosols cleared. This was reported as the normalised and absolute aerosol concentration decay over time.

Intervention tests included the following:

- Effect of installation of portable air cleaners in the patient room vs usual HVAC on aerosols within and external to the room
- Effect of patient room door open vs door closed on aerosols outside the room
- Effect of installation of variable number of portable air cleaners in the corridor on aerosols in the corridor
- Effect of installation of different barriers to enclose the nurse’s station on aerosols in within the nurses’ station

For example, the time taken to clear the patient room and the nurses’ station from aerosols under existing HVAC settings at an air change rate of 12 times per hour was compared to the time taken when air cleaners were placed in the patient room. The air cleaners used were domestic appliances (Samsung™ AX5500K) equipped with H13 HEPA filters capable of filtering 99.97% of particles at a Clean Air Delivery Rate of 467 m^3^/hr on the highest fan speed setting. Based on laboratory testing, for the size of the hospital room and the capacity of the air cleaner selected, two air cleaners were used and were placed along the patient’s bedside and foot of the bed. The time taken to clear the aerosols from the patient room and the nurses’ station opposite was measured when the patient room had the door open and when the door was closed, for comparison. The impact of additional protections for the nurses’ station included when the nurses’ station was protected by an air cleaner “barrier” or a plastic Zipwall™. An air cleaner “barrier” consisted of three air cleaners lined up 1m apart directly in front of the desk. The ZipWall ™ is a clear plastic barrier that was erected in front of the desk, sealed at the ceiling and walls on either side, with a magnetised self-closing door for people to enter and exit the space. Measurements of aerosol clearance were also taken in the corridor of the ward which was approximately 50m long, and 2m wide. The corridor was flooded with smoke and the rates of clearance of aerosols were compared with different numbers of air cleaners positioned along its length to identify the optimum number of air cleaners for that space.

## Results

At baseline, the air in the patient room had negligible demonstrable aerosols, and after smoke flooding was applied, the entire room was rapidly and visibly filled with smoke. During the tests with the patient room door closed, within the first minute after the room was flooded with smoke, smoke was observed to escape under the gap at the bottom of the door and move along the corridor towards the return air vent at the entrance of the ward. When the door to the patient room was opened, smoke was observed to immediately move out to the corridor and travel along the corridor to the return air vent.

Using measurements within the patient room, at usual HVAC settings, and with a closed door it took 16 minutes for the aerosols in the patient room to clear back down to 1% of the baseline maximum measurable by the instruments. The concentration of aerosols concurrently reaching the nurses’ station even when the patient room door was closed were high and decreased at the same rate as aerosols inside the room. The relative amount of aerosols in the nurses’ station compared to the patient room was highly variable representing 25-50% of the room’s concentration with the door closed using two air cleaners. When two air cleaners were placed in the patient room with the door closed or open, the room cleared of 99% aerosols in 5.5 minutes – a 67% reduction compared with no air cleaners – and the smoke at the nurses’ station cleared even more quickly, at under 3 minutes (Figure 1). Having the bathroom door open with exhaust fan running made a negligible difference to the clearance time (∼50 seconds, and within the variability of the observations).

**Figure 1:**
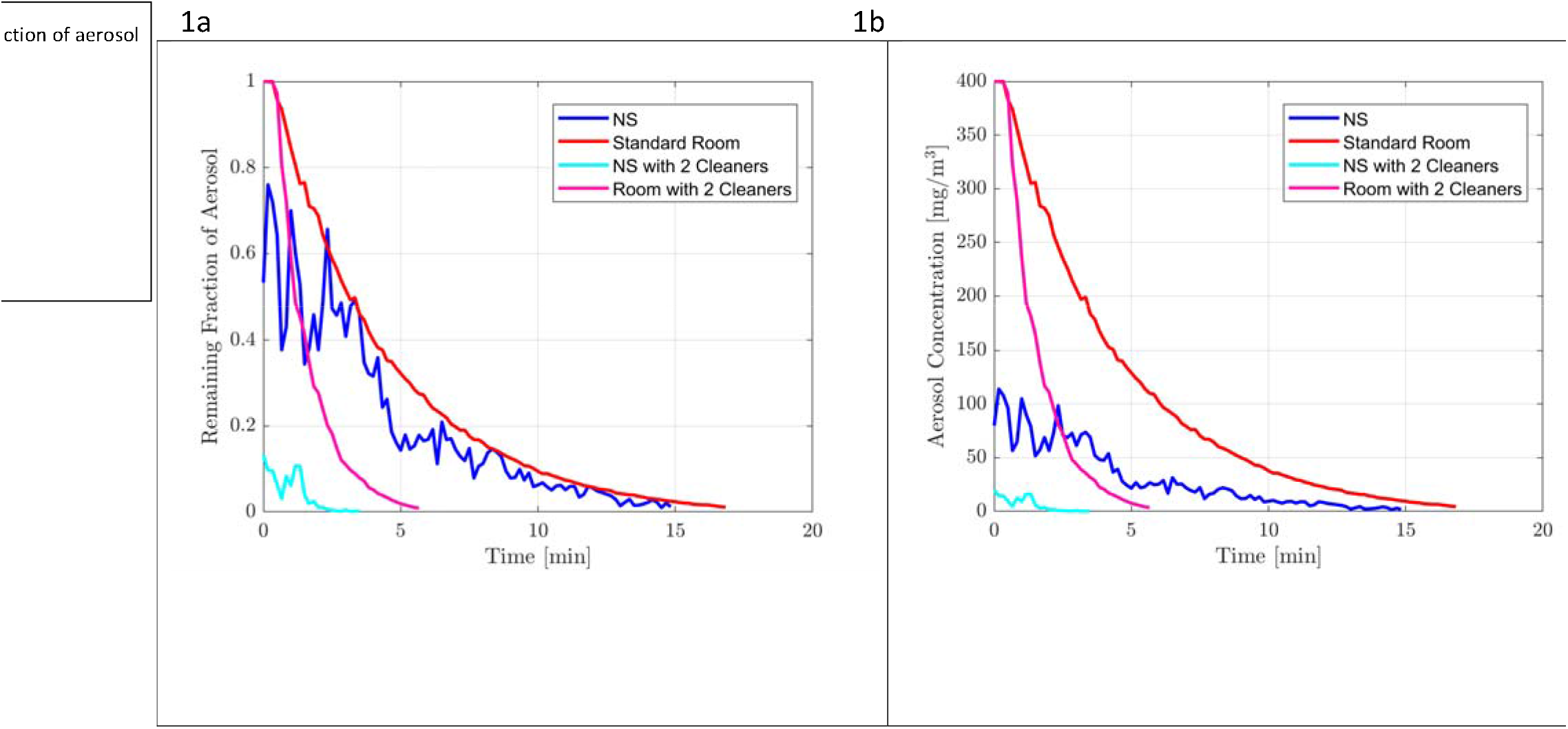
Comparison of the effect of no air cleaners (just usual heating ventilation and air conditioning) versus 2 air cleaners on aerosol clearance and transmission of aerosols. Measurement taken within a standard patient room and at the nurses’ station (NS) with the door closed. 1a shows the values normalized to the saturation value of the sensor 1b shows the measured value

When the door to the patient room was opened, high levels of aerosols had crossed the corridor and entered the nurses’ station at baseline measurement, as the air cleaners were used in the patient room the aerosols cleared from the nurses’ station over 4 minutes. When the door to the patient room was closed, and air cleaners were used in the patient room, very low levels of aerosol were detectable at the nurses’ station at baseline and 99% of these cleared within approximately 2 minutes (Figure 2)

**Figure 2:**
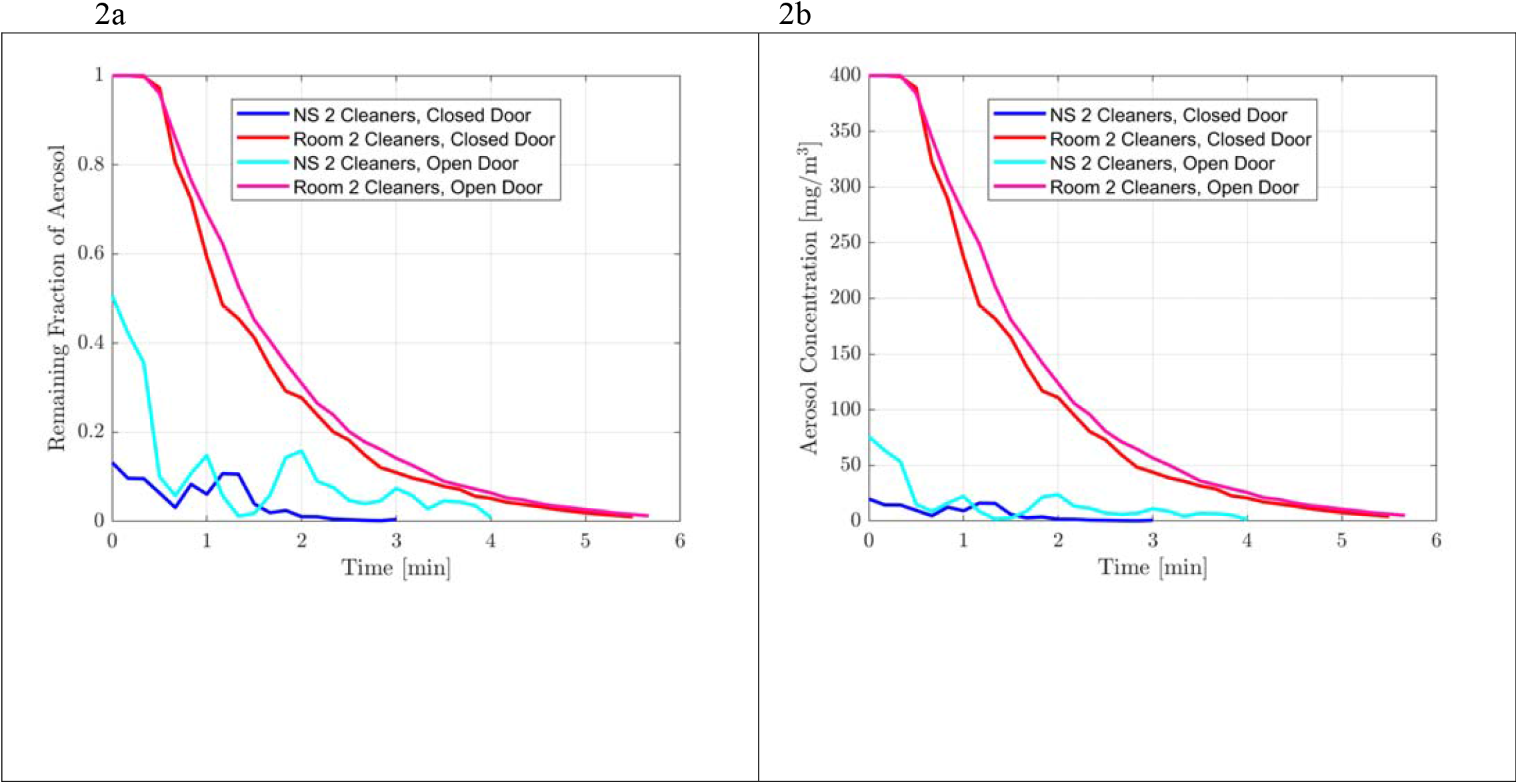
Comparison of the effect of open vs closed door to patient room on aerosol clearance and transmission of aerosols. Measurement taken within a standard patient room and at the nurses’ station (NS) with 2 air cleaners in the patient room. 2a shows the values normalized to the saturation value of the sensor 2b shows the measured value.

Two different barriers were tested to determine the most effective way to provide protection to the nurses’ station. When the air cleaner barrier was used, negligible smoke particles from the patient room were measured at the nurses’ station (Figure 3). Similarly the ZipWall™ substantially reduced the ability of the aerosolized smoke particles from entering the nurses’ station (Figure 3).

**Figure 3:**
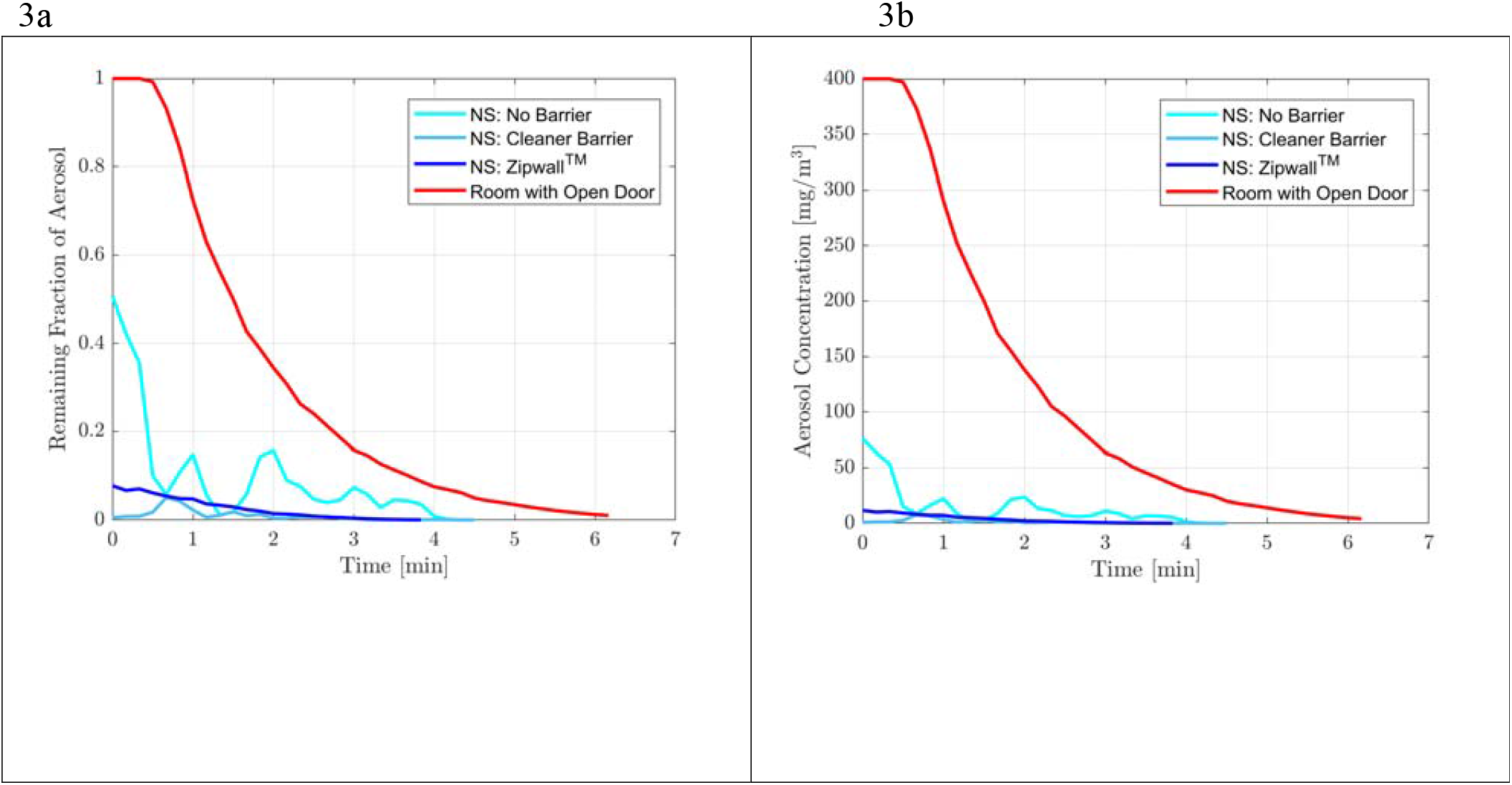
Comparison of different interventions at the nurses’ station on aerosol clearance and transmission of aerosols. Measurement taken within a standard patient room and at the nurses’ station (NS) with 2 air cleaners in the patient room. 3a on left shows the values normalized to the saturation value of the sensor and 3b on the right shows the measured value. Zipwall ™ = plastic sheeting in front of main desk, taped at roof, floor and sides with magnetically closing doorway. Air cleaner barrier = 3 air cleaners in front of desk

When the corridor was flooded with smoke, between 4 and 12 air cleaners were evenly spaced along its length. Adding extra air cleaners did not increase the clearance much beyond that which was achieved with 8 air cleaners, which achieved 99% clearance in 5 minutes (Figure 4).

**Figure 4:**
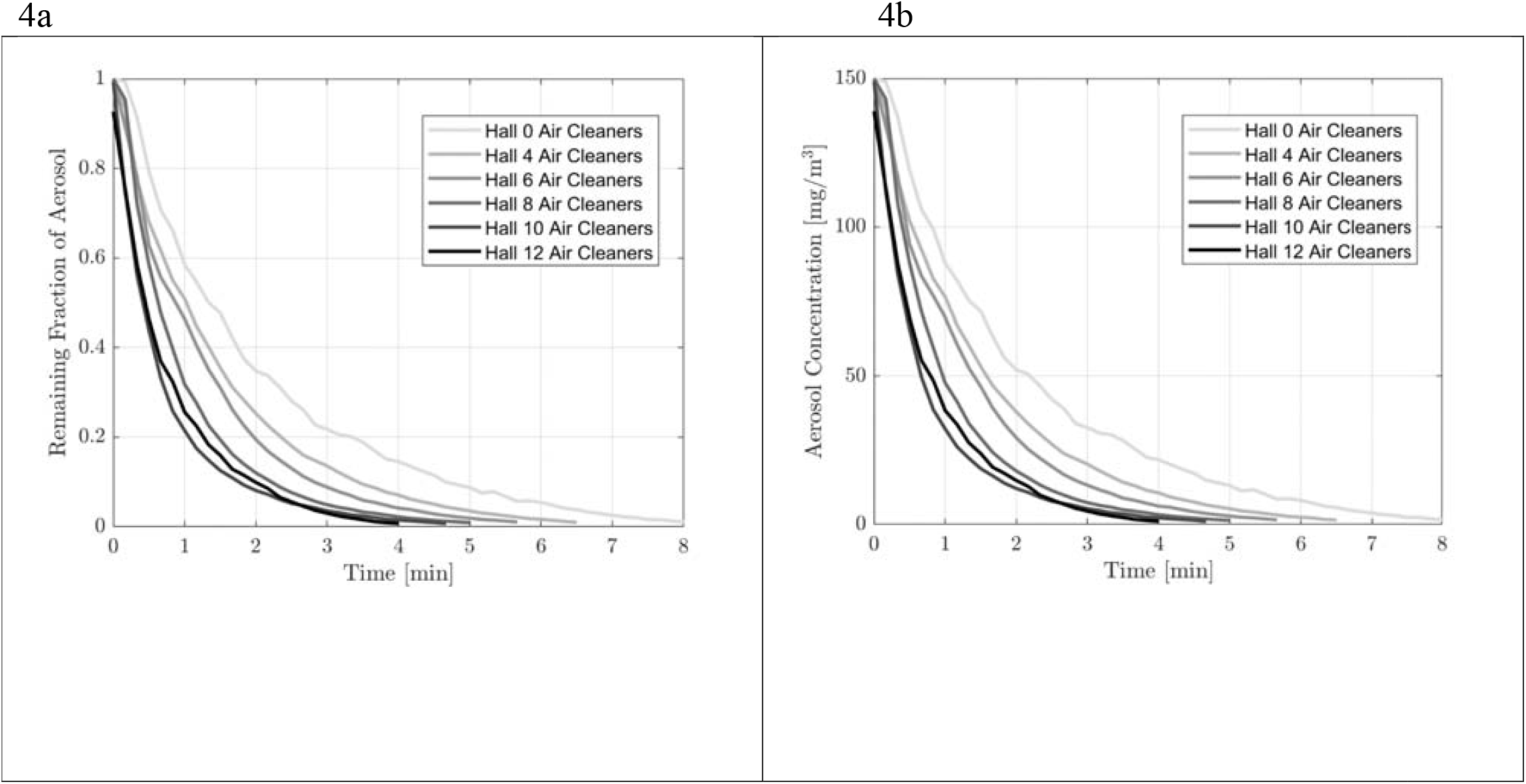
Rate of clearance of aerosolized smoke particles in the corridor with differing numbers of air cleaners in that corridor. 4a on left shows the values normalized to the saturation value of the sensor and 4b on the right shows the measured value.

## Discussion

This study demonstrated that the existing ward HVAC system alone was quite poor at clearing a patient room of aerosols, and suggests that commercially available air cleaners may have a role in clearing aerosolised particles that may contain respiratory viruses, such as SARS-CoV-2, in clinical environments. We found that the actual 99% clearance of aerosols from the air in patient rooms using existing HVAC alone at 12 air exchanges/hour was quite slow. Aside from isolation rooms, HVAC systems in hospitals and other places are designed for comfort rather than infection control. It may be worth noting that air exchange rates of 2 would not be uncommon in an office environment. In an Australian hospital environment 6 air exchanges/hour is the standard, and 10 has been suggested for wards managing COVID-19 patients. These data demonstrate that the relationship between reported air exchanges and actual aerosol clearance from rooms is not predictable, probably because it does not account for flow recirculation regions or other air flow anomalies. Theoretically, to clean 99% of the air of a well-mixed room in under 10 minutes would require the same air exchange equivalent to air exchanges of 30/hour.^31^ For context, typical operating theatres have 20 air exchanges/hour,^32, 33^and there is advice to wait for 5 full air exchanges (18 minutes for 99.3% clearance) until staff can enter without airborne precautions where necessary.^34^ Air exchanges greater than 30 are impossible to achieve with the typical hospital ward HVAC systems alone, but not difficult with air cleaners.

Our data suggest that any air that enters a patient room needs to travel to reach a return air vent. In some cases, return air vents are placed in the patient room or in the ensuite bathroom. In the ward we studied, the position of the return air vent meant that air was leaving the patient room and travelling along the corridor. It is important to understand where and how air exits a patient’s room so that the path of least resistance for air to travel from the patient to the return air vent can be considered when patients are placed on a ward. On the ward studied, the preference would now be to utilise rooms close to that air return for higher risk patients (e.g.; earlier in their illness, actively coughing) where possible. These findings also confirmed that the doors to the patient rooms should be kept closed wherever possible to protect areas outside the recognised patient zone, appreciating that air and the aerosols carried in it may still travel. This study showed that air cleaners in the patient room limit how much aerosol is likely to escape into the corridor or nurses’ station, which likely provides protection for staff. It also showed that air cleaners placed along the corridor should be considered (in addition to cleaners in the patient room) if the ward was filled with patients infected with SARS-CoV-2, again to help clear the air of potentially infected aerosols that may have escaped from the patient’s room. Finally a physical barrier, such as a ZipWall™, may provide additional protection for the staff if they spend time at a workstation.

Importantly this work used glycerine based aerosol as a proxy for respiratory aerosols that contain live virus. Extrapolation must be made from other studies in which viral RNA or culturable virus has been identified in aerosols from clinical spaces caring for people with SARS-CoV-2 infection.^26, 27, 29, 35^ Nevertheless, it demonstrates that aerosols of the same size as respiratory particles can travel long distances. Viral transmission via aerosols is likely to be very variable, both between patients, and at different times in a given patient’s infection course and may be impacted by other factors such as relative humidity and temperature.^36^ We hypothesise that if multiple patients are in a given confined space, each producing some respiratory particles, then the density of infectious aerosols in the air may become high enough to put staff at increased risk of acquisition via aerosols, even in corridors or nurses’ stations of dedicated COVID-19 wards. Given that the air cleaners remove aerosols from the air before it leaves the patient room, it is likely that they would help protect staff inside and outside the patient room. These air cleaners are a relatively low cost, readily implementable mitigation strategy that should be considered in clinical spaces. These findings may also have implications outside healthcare institutions that warrant further investigation.

## Conclusion

Despite exceeding recommended air exchange rates for a hospital, the HVAC alone did not effectively remove aerosols from the clinical space in a timely matter. In addition, depending on the location of the return air duct, the existing HVAC system promoted the dispersal of aerosols beyond the patient room. However we were able to demonstrate that relatively low-cost air cleaners could dramatically increase the clearance rate of aerosols. Air in clinical spaces does travel, and with it any respiratory aerosols containing potentially infectious virus. An understanding of directional airflow is important to help limit the risk to staff, and will likely be different in different spaces. Based on the findings of this work, the hospital studied has adopted air cleaners for use in rooms of patients with suspected or confirmed COVID-19, with appropriate policies, procedures and training to ensure their safe and effective use.

## Data Availability

The data are held by the researchers at the University of Melbourne

## Financial support

The work was funded by The Royal Melbourne Hospital

## Potential conflicts of interest

All authors report no conflicts of interest relevant to this article

## Supplementary figures

**Figure S1:**
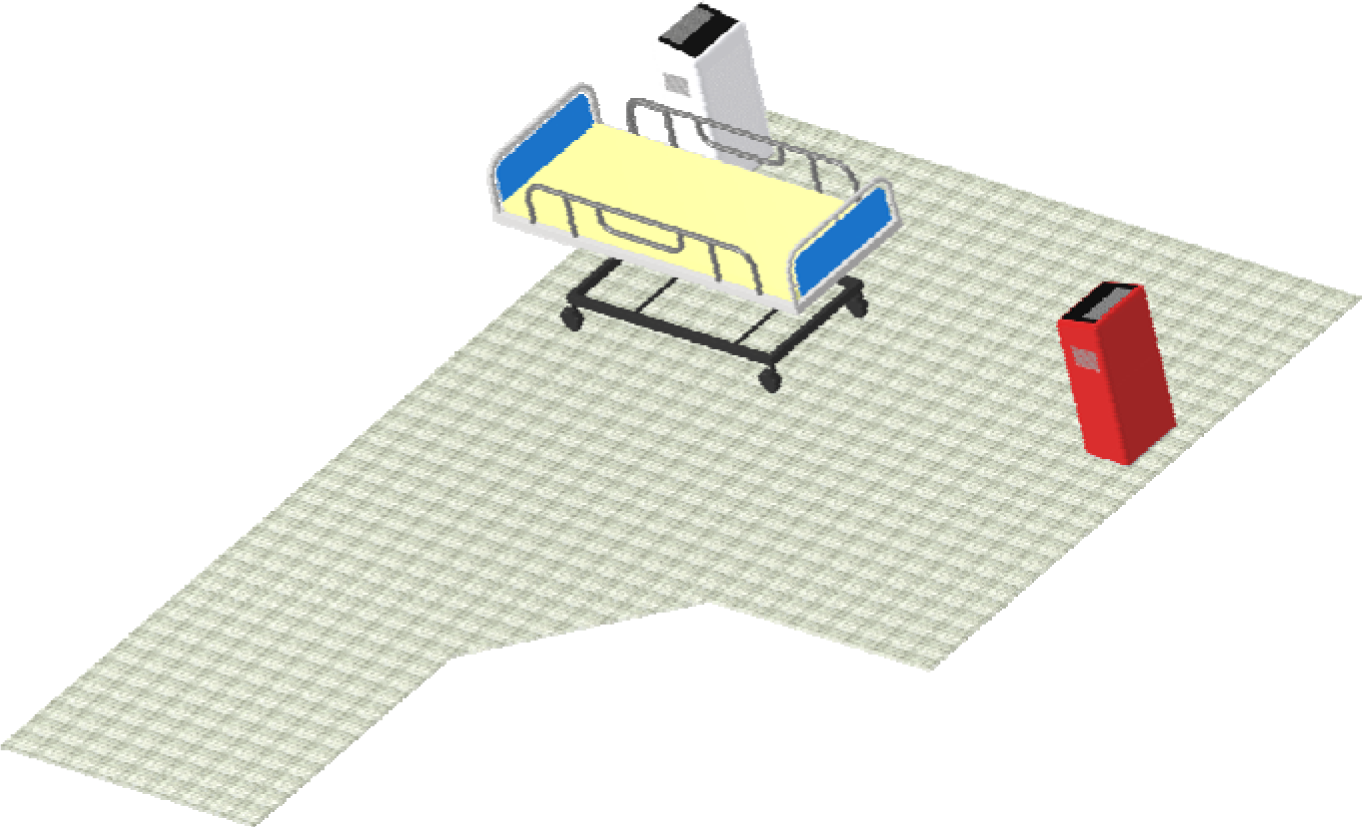
Isometric view of a computer rendering of the single patient room. Room entry would be from the lower left hand corner of the image. The white air cleaner (or portable HEPA filter) is the one on the patient bedside and the red air clener (portable HEPA filter) is flush with the wall

**Figure S2:**
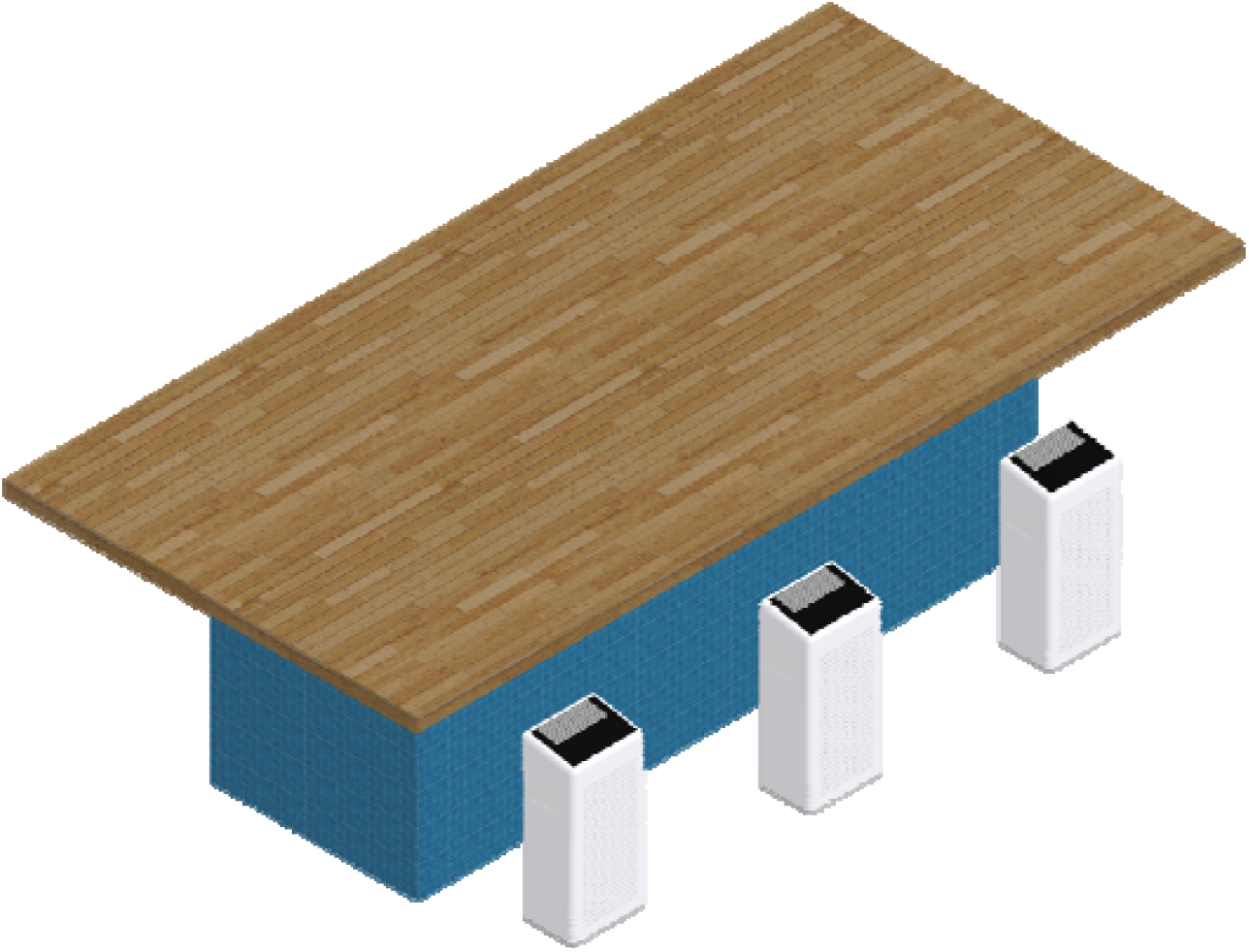
Isometric view of a computer rendering of the air cleaner barrier in front of the nurses’ station table.

**Figure S3:**
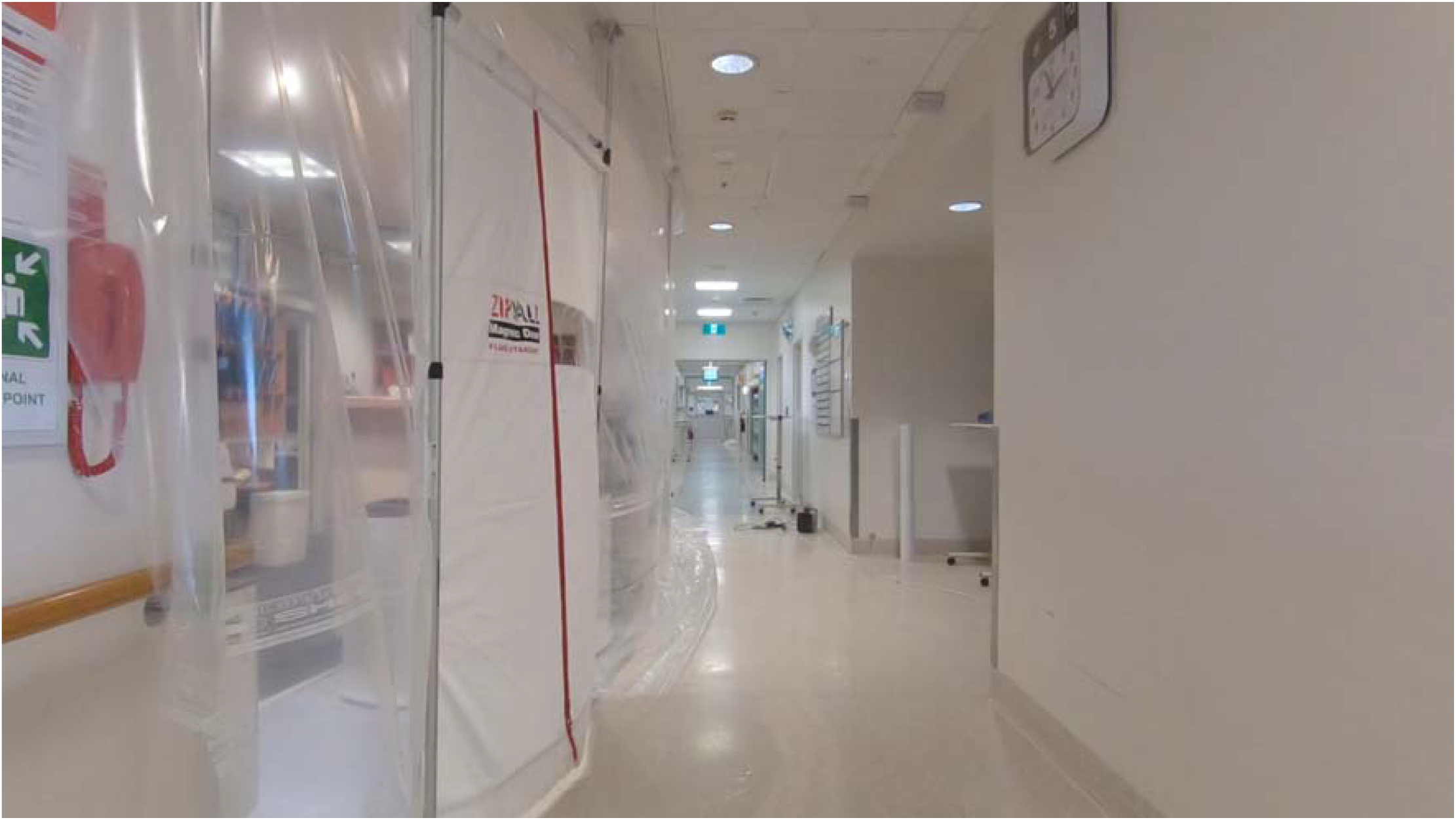
Photo of installed ZipWall™ at nurses’ station

**Figure S4:**
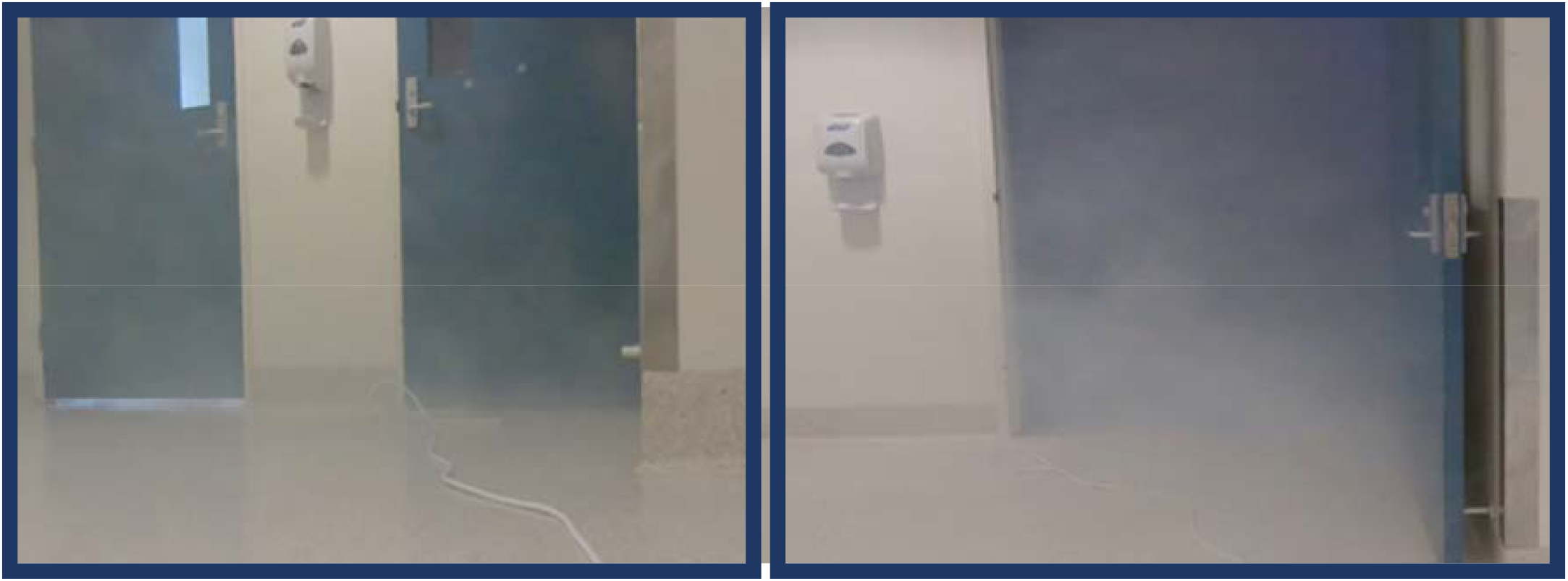
Comparison of smoke exiting the patient room with the door closed (left) and open (right). Note that the figures were taken at approximately the same time after the room was flooded with smoke.

**Figure S5:**
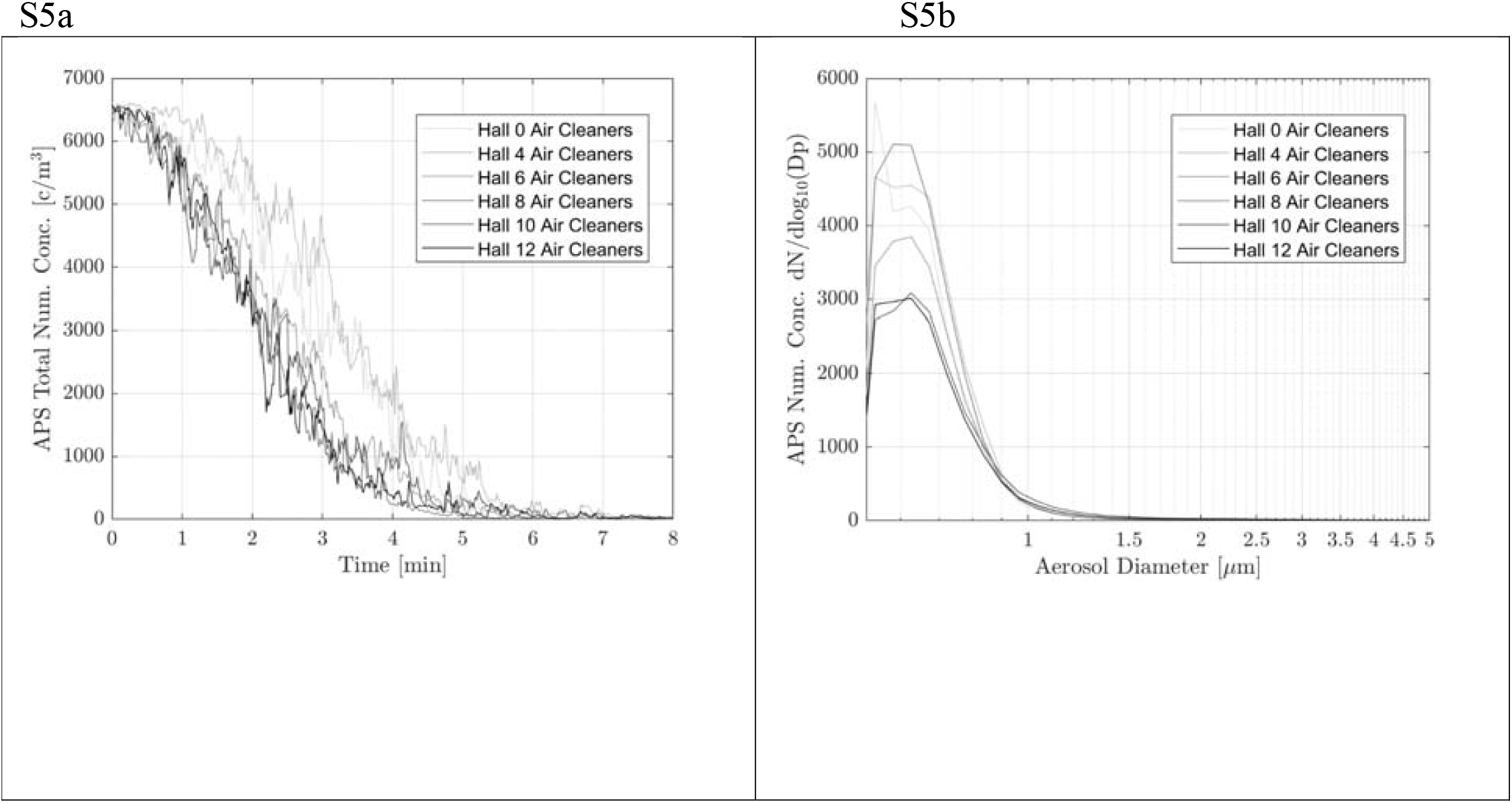
Aerodynamic Particle Sizer™ (APS) particle number size distribution observations for the variable number of air cleaners during corridor clearance experiment. S5a is the same as 4b but showing the APS observed number concentrations S5b shows the APS observed size number concentrations.

## Notes

### Competing Interest Statement

The authors have declared no competing interest.

### Author Declarations

The Royal Melbourne Hospital human research ethics committee reviewed the manuscript and determined that it was exempt from requiring ethics approval

